# Differential asthma risk following respiratory infection in children from three minority populations

**DOI:** 10.1101/19011528

**Authors:** Eric M. Wohlford, Luisa N. Borrell, Jennifer R. Elhawary, Brian Plotkin, Sam S. Oh, Thomas J. Nuckton, Celeste Eng, Sandra Salazar, Michael A. LeNoir, Kelley Meade, Harold J. Farber, Denise Serebrisky, Emerita Brigino-Buenaventura, William Rodriguez-Cintron, Rajesh Kumar, Shannon Thyne, Max A. Seibold, José R. Rodriguez-Santana, Esteban G. Burchard

## Abstract

**Rationale:** Severe early-life respiratory illnesses, particularly those caused by respiratory syncytial virus (RSV) and human rhinovirus (HRV), are strongly associated with the development of asthma in children. Puerto Rican children in particular have a strikingly high asthma burden, but prior studies of the potential associations between early-life respiratory illnesses and asthma in Puerto Rican and other minority populations have been limited.

**Objectives:** We sought to determine whether early-life respiratory illness was associated with asthma in Puerto Rican children relative to other minority children.

**Methods:** Using a logistic regression analysis, we examined the association between early-life respiratory illnesses (report of upper respiratory infection (URI), pneumonia, bronchitis, and bronchiolitis/RSV) within the first two years of life and physician-diagnosed asthma after the age of two in a large cohort of minority children.

**Measurements and Main Results:** Early-life respiratory illnesses were associated with greater asthma risk in Puerto Ricans relative to other racial/ethnic minority populations. Specifically, in Puerto Ricans, the odds was 6.15 (95% CI: 4.21-9.05) if the child reported at least one of the following respiratory illness: URI, pneumonia, bronchitis or bronchiolitis. The odds were also higher in Puerto Ricans when considering these conditions separately.

**Conclusions:** We observe population-specific associations between early-life respiratory illnesses and asthma, which was especially significant in Puerto Ricans. Taken together with the known high burden of RSV in Puerto Rico, our results may help explain the high burden of asthma in Puerto Ricans.

## Introduction

Asthma is the most common chronic disease in children[1, 2], with genetic, environmental, and infectious risk factors.[3-5] Though the global burden of asthma is increasing, certain racial/ethnic and geographic populations are at especially high risk. Puerto Ricans are among the most severely affected populations in the world.[4] Approximately 36.5% of Puerto Ricans report they currently or previously had asthma, compared to only 13.0% of African Americans, 12.1% of European Americans, and 7.5% of Mexican Americans.[6] These striking differences extend to asthma morbidity and mortality, which are 2.4- and 4-fold higher in Puerto Ricans compared to whites, respectively.[6, 7]

Several epidemiological studies have established a strong association between the development of childhood asthma or recurrent wheeze with exposure to severe, early-life respiratory illnesses across populations.[8-18] Associations with asthma were strongest for infections caused by respiratory syncytial virus (RSV)[8, 11, 14-18] and human rhinovirus (HRV).[12-14] Both RSV and HRV are known to cause bronchiolitis, which can be a severe respiratory infection in children and is linked to later asthma development.[16, 19] These viruses have a complex interaction between genetics and environmental exposures in determining risks for asthma and related outcomes.[20, 21] Additionally, Puerto Rico has an RSV season that is year-round whereas the mainland United States only reports a 20-week season (**Fig 1**).[22, 23]

**Fig 1.**
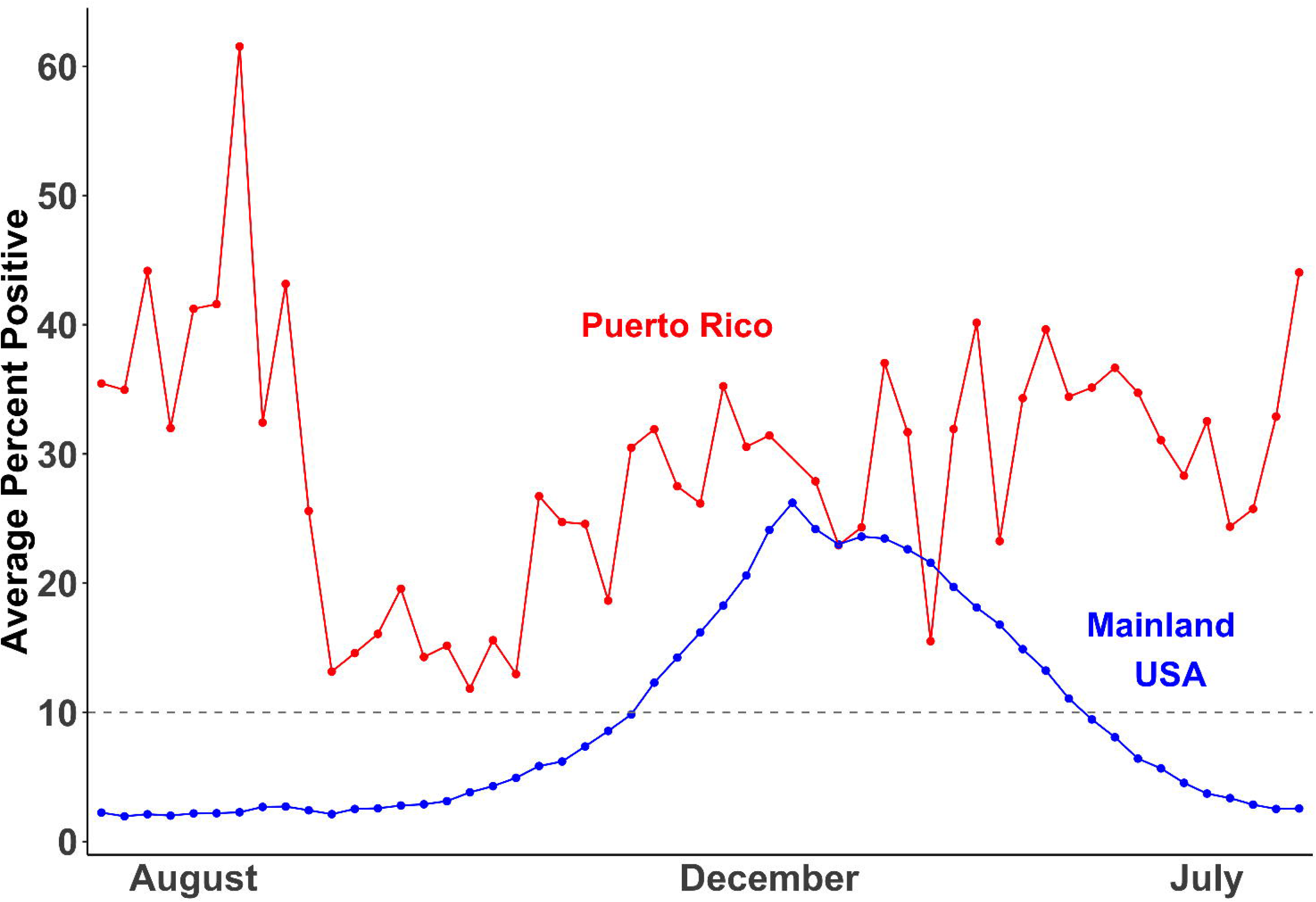
Respiratory syncytial virus (RSV) season in Puerto Rico (red) and the mainland United States (blue). RSV season begins when 10% or more of RSV tests are positive.

It is unclear at this time if differential responses to early-life respiratory illnesses contribute to the striking asthma disparities seen across minority populations. It is possible that genetic predisposition, environmental influences, and early-life respiratory illness work together to increase asthma susceptibility in high-risk populations. Our aim was to investigate the association of early-life respiratory illnesses with asthma susceptibility seen in our large and well-phenotyped cohort of minority children. In light of disparate disease prevalence,[6, 22, 23] we examined this association in each population (Puerto Ricans, African Americans and Mexican Americans), separately and combined.

## Methods

### Study population

The analysis examined participants recruited through two parent studies: the Genes-Environments and Admixture in Latino Americans (GALA II) study and the Study of African Americans, Asthma, Genes and Environments (SAGE), described in detail elsewhere.[24-26] Briefly, GALA II and SAGE are parallel case-control studies of asthma conducted between 2006 to 2014 in Latino (Mexican American and Puerto Rican) and African American children aged 8-21, respectively. SAGE participants were recruited from the San Francisco Bay Area and GALA II participants were recruited from across the continental United States (Chicago, Houston, New York City, and the San Francisco Bay Area) and Puerto Rico. Questionnaires were administered to adult participants and parents of minors. All participants provided consent to being in the study. Consent was obtained from all adult participants and parent/legal guardians of minor participants. All participating institutions obtained the appropriate approvals from their Institutional Review Boards.

Out of a total of 6,023, participants with asthma onset prior to the age of two (n=1,450) and those individuals without complete illness and covariate data (n=2,301) were excluded from the study. Those diagnosed with asthma before age two were initially excluded from analysis to better delineate any causal association between early-life respiratory illness before age two and subsequent diagnosis of asthma. The exclusion criteria yielded an analytical sample of 2,824 subjects, including both cases (n=1,091) and controls (n=1,733).

### Outcomes

The outcome of our study was asthma diagnosed after the age of two by a physician.[24] Eligible control subjects had no reported history of asthma, lung disease, or chronic illness, and no reported symptoms of coughing, wheezing, or shortness of breath in the two years before enrollment.

### Exposure

The exposure was self-reported early-life respiratory illnesses within the first two years of life. Early-life respiratory illnesses included upper respiratory infection (URI), pneumonia, bronchitis, and bronchiolitis/RSV, which were analyzed separately. In addition, the report of at least one of the four previous illnesses was used to categorize children with “any respiratory illness” and none otherwise.

### Covariates

Consistent with previous studies,[27, 28] we considered factors known to be associated with early-life respiratory illnesses and with asthma. These included sex, underweight at birth (yes/no), maternal smoking during pregnancy (yes/no), whether the subject was breastfed (yes/no), number of older siblings (none, one, two or more), socioeconomic status (SES; low, medium, high; described further in **Text A in S1 File**), and recruitment region; the latter of which was not used in the African American models because all the participants were recruited from the same region. Global African and European ancestry estimates (described further in **Text B in S1 File**) were additional covariates used in our combined model to adjust for the underlying substructure of populations in the dataset.

### Statistical Analysis

Descriptive statistics were calculated for the overall study population and for each minority population separately. Logistic regression was used to study the association between physician-diagnosed asthma and self-reported respiratory illnesses in the first two years of life. To account for the difference in asthma prevalence[6] and the fact that ethnicity acts as an effect modifier on the association, we also performed association tests stratified by population. The statistical programming language R version 3.5.1 was used to perform all analyses.

### Data availability

Due to the sensitive nature of the data collected surrounding issues of race, socioeconomic status, and ancestry, data are available upon request. Access to the limited dataset used in this study can be arranged by contacting Dr. Esteban Burchard. Of note, any data provided will be stripped of specific subject identifiers that could be used to identify a specific child and/or his/her residence. All source code for this analysis can be accessed on our github page https://github.com/asthmacollaboratory/asthma-after-RI.

## Results

Descriptive characteristics of our final study population are presented in **Table 1**. When compared with children without asthma, those with asthma were more likely to be male, to have a mother who smoked during pregnancy, and have lower SES. These distributions were observed in our sample with a few exceptions. For instance, while Puerto Rican and Mexican American children without asthma were more likely to be breastfed than their counterparts with asthma, the opposite was true for African Americans. Additionally, we found that Puerto Ricans were 4- to 9-fold more likely to report an RSV infection or bronchiolitis in the first two years of life than Mexican Americans and African Americans, respectively (**Fig 2**).

**Table 1.** Descriptive statistics of the final study population (N = 2,824).

|  | Puerto Rican |  |  |  | Mexican American |  |  |  | African American |  |  |  | Combined |  |  |  |
| --- | --- | --- | --- | --- | --- | --- | --- | --- | --- | --- | --- | --- | --- | --- | --- | --- |
|  | Case |  | Control |  | Case |  | Control |  | Case |  | Control |  | Case |  | Control |  |
|  | N | % | N | % | N | % | N | % | N | % | N | % | N | % | N | % |
| Number of subjects | 284 | 100 | 783 | 100 | 375 | 100 | 504 | 100 | 432 | 100 | 446 | 100 | 1091 | 100 | 1733 | 100 |
| Males | 150 | 52.8 | 361 | 46.1 | 212 | 56.5 | 210 | 41.7 | 218 | 50.5 | 187 | 41.9 | 580 | 53.2 | 758 | 43.7 |
| Underweight at birth | 32 | 11.3 | 61 | 7.8 | 24 | 6.4 | 35 | 6.9 | 36 | 8.3 | 45 | 10.1 | 92 | 8.4 | 141 | 8.1 |
| In-utero smoke exposure | 14 | 4.9 | 40 | 5.1 | 14 | 3.7 | 9 | 1.8 | 78 | 18.1 | 54 | 12.1 | 106 | 9.7 | 103 | 5.9 |
| Breastfed | 141 | 49.6 | 432 | 55.2 | 285 | 76 | 401 | 79.6 | 262 | 60.6 | 244 | 54.7 | 688 | 63.1 | 1077 | 62.1 |
| Number of older siblings |  |  |  |  |  |  |  |  |  |  |  |  |  |  |  |  |
| 0 | 74 | 26.1 | 200 | 25.5 | 132 | 35.2 | 195 | 38.7 | 247 | 57.2 | 208 | 46.6 | 453 | 41.5 | 603 | 34.8 |
| 1 | 92 | 32.4 | 274 | 35 | 138 | 36.8 | 176 | 34.9 | 101 | 23.4 | 132 | 29.6 | 331 | 30.3 | 582 | 33.6 |
| 2 or more | 118 | 41.5 | 309 | 39.5 | 105 | 28 | 133 | 26.4 | 84 | 19.4 | 106 | 23.8 | 307 | 28.1 | 548 | 31.6 |
| Socioeconomic status* |  |  |  |  |  |  |  |  |  |  |  |  |  |  |  |  |
| high | 89 | 31.3 | 271 | 34.6 | 114 | 30.4 | 116 | 23 | 163 | 37.7 | 160 | 35.9 | 366 | 33.5 | 547 | 31.6 |
| medium | 51 | 18 | 136 | 17.4 | 79 | 21.1 | 84 | 16.7 | 113 | 26.2 | 137 | 30.7 | 243 | 22.3 | 357 | 20.6 |
| low | 144 | 50.7 | 376 | 48 | 182 | 48.5 | 304 | 60.3 | 156 | 36.1 | 149 | 33.4 | 482 | 44.2 | 829 | 47.8 |
| Recruitment region |  |  |  |  |  |  |  |  |  |  |  |  |  |  |  |  |
| Chicago | 18 | 6.3 | 23 | 2.9 | 137 | 36.5 | 180 | 35.7 | - | - | - | - | 155 | 14.2 | 203 | 11.7 |
| Houston | 1 | 0.4 | - | - | 96 | 25.6 | 91 | 18.1 | - | - | - | - | 97 | 8.9 | 91 | 5.3 |
| New York | 27 | 9.5 | 34 | 4.3 | 20 | 5.3 | 64 | 12.7 | - | - | - | - | 47 | 4.3 | 98 | 5.7 |
| San Francisco Bay Area | 1 | 0.4 | 1 | 0.1 | 122 | 32.5 | 169 | 33.5 | 432 | 100 | 446 | 100 | 555 | 50.9 | 616 | 35.5 |
| Puerto Rico | 237 | 83.5 | 725 | 92.6 | - | - | - | - | - | - | - | - | 237 | 21.7 | 725 | 41.8 |
| URI | 57 | 20.1 | 35 | 4.5 | 24 | 6.4 | 27 | 5.4 | 67 | 15.5 | 16 | 3.6 | 148 | 13.6 | 78 | 4.5 |
| Pneumonia | 13 | 4.6 | 5 | 0.6 | 20 | 5.3 | 11 | 2.2 | 24 | 5.6 | 14 | 3.1 | 57 | 5.2 | 30 | 1.7 |
| Bronchitis | 36 | 12.7 | 10 | 1.3 | 37 | 9.9 | 10 | 2 | 16 | 3.7 | 6 | 1.3 | 89 | 8.2 | 26 | 1.5 |
| Bronchiolitis/RSV | 31 | 10.9 | 14 | 1.8 | 6 | 1.6 | 4 | 0.8 | 6 | 1.4 | 2 | 0.4 | 43 | 3.9 | 20 | 1.2 |
| Any Listed | 89 | 31.3 | 54 | 6.9 | 75 | 20 | 46 | 9.1 | 98 | 22.7 | 31 | 7 | 262 | 24 | 131 | 7.6 |

\*Socioeconomic status was derived from a combination of mother's education level, health insurance status, and household income weighted by region, see **Text A in S1 File** for more information.

**Fig 2.**
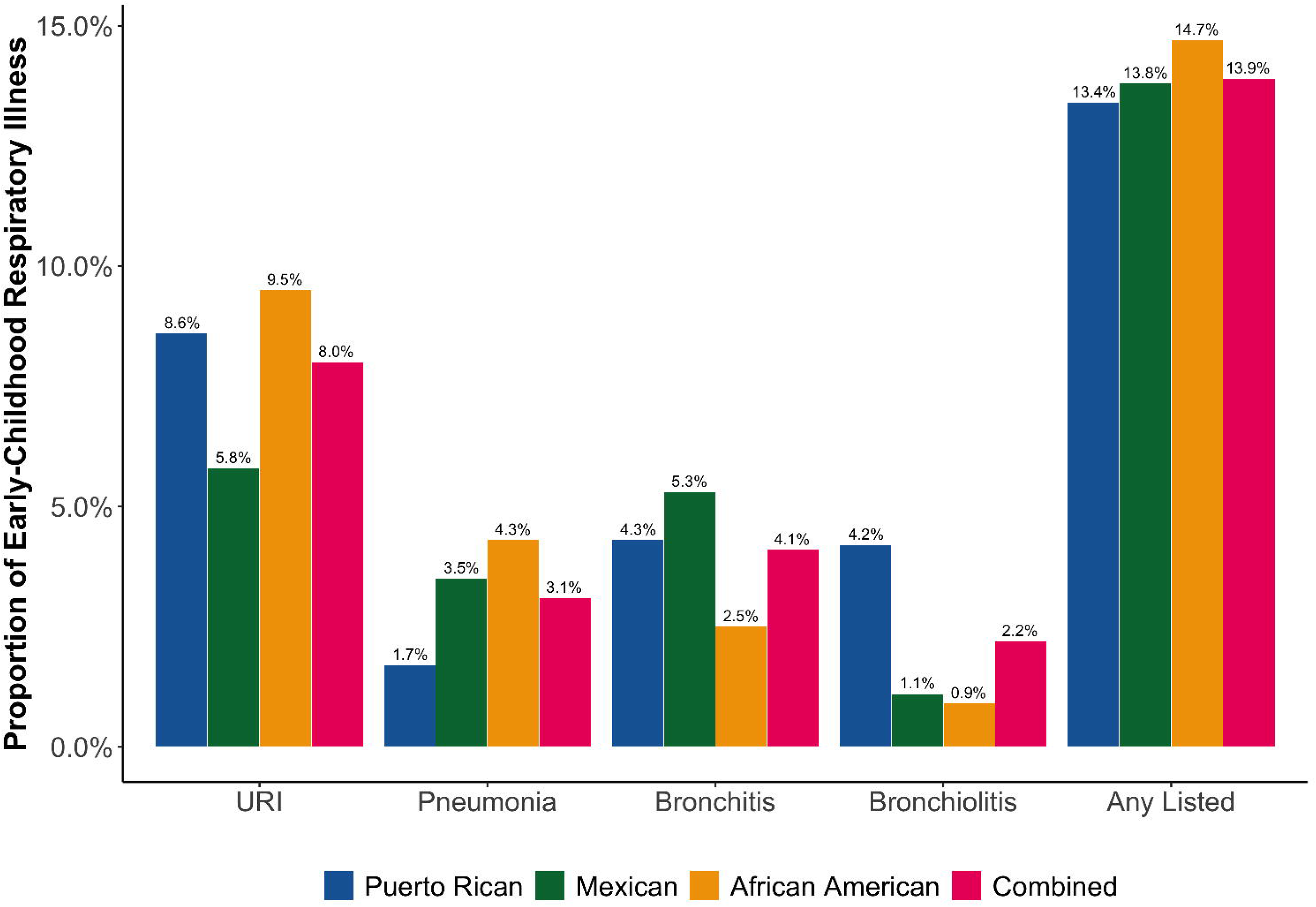
Population-specific proportions of selected respiratory illnesses during the first 2 years of life for our study population.

In the population-specific stratified analyses, we also observed that the odds of asthma in Puerto Ricans was 5.25 (95% CI: 3.34-8.37) for URI, 7.23 (95% CI: 2.66-23) for pneumonia, 13 (95% CI: 6.51-28.2) for bronchitis, 7.27 (95% CI: 3.83-14.5) for bronchiolitis/RSV, and 6.15 (95% CI: 4.21-9.05) if the participant reported having at least one early-life respiratory illness (**Table 2, Fig 3**). Comparatively, the odds of asthma in Mexican Americans and African Americans respectively was 2.17 (95% CI: 1.12-4.24) and 4.77 (95% CI: 2.76-8.71) for URI, 2.05 (95% CI: 0.956-4.61) and 1.9 (95% CI: 0.967-3.86) for pneumonia, 4.78 (95% CI: 2.39-10.4) and 2.7 (95% CI: 1.08-7.71) for bronchitis, 2.01 (95% CI: 0.552-8.15) and 2.9 (95% CI: 0.635-20.4) for bronchiolitis/RSV, and 3.07 (95% CI: 1.99-4.79) and 3.91 (95% CI: 2.55-6.12) if the participant reported having at least one early-life respiratory illness (**Table 2, Fig 3**). In our combined analysis, we observed that the odds of asthma was 4.31 (95% CI: 3.15-5.96) for URI, 2.66 (95% CI: 1.67-4.3) for pneumonia, 7.04 (95% CI: 4.44-11.6) for bronchitis, 5.82 (95% CI: 3.26-10.8) for bronchiolitis/RSV, and 4.5 (95% CI: 3.52-5.78) if the participant reported having at least one early-life respiratory illness (**Table 2, Fig 3**).

**Table 2.**
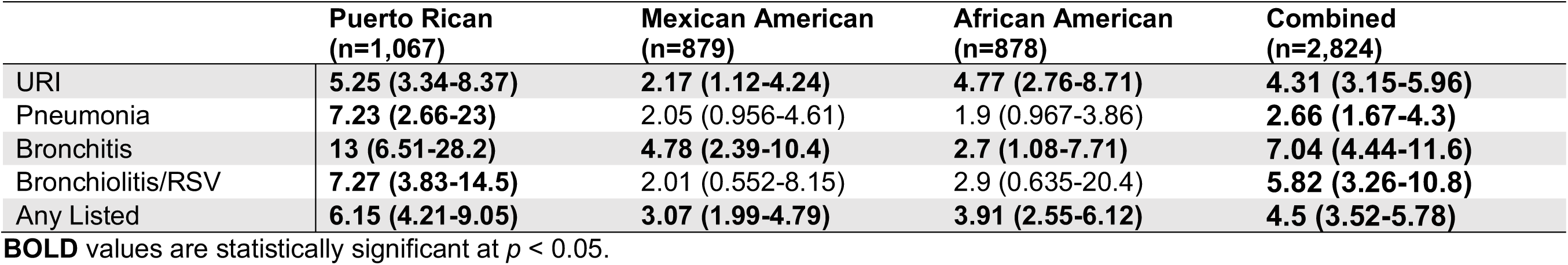
Odds ratios and confidence intervals from the population-specific and combined association analysis between respiratory illnesses in the first two years of life and physician-diagnosed asthma after the age of two in the three main populations in GALA II and SAGE: 2006-2014.

**Fig 3.**
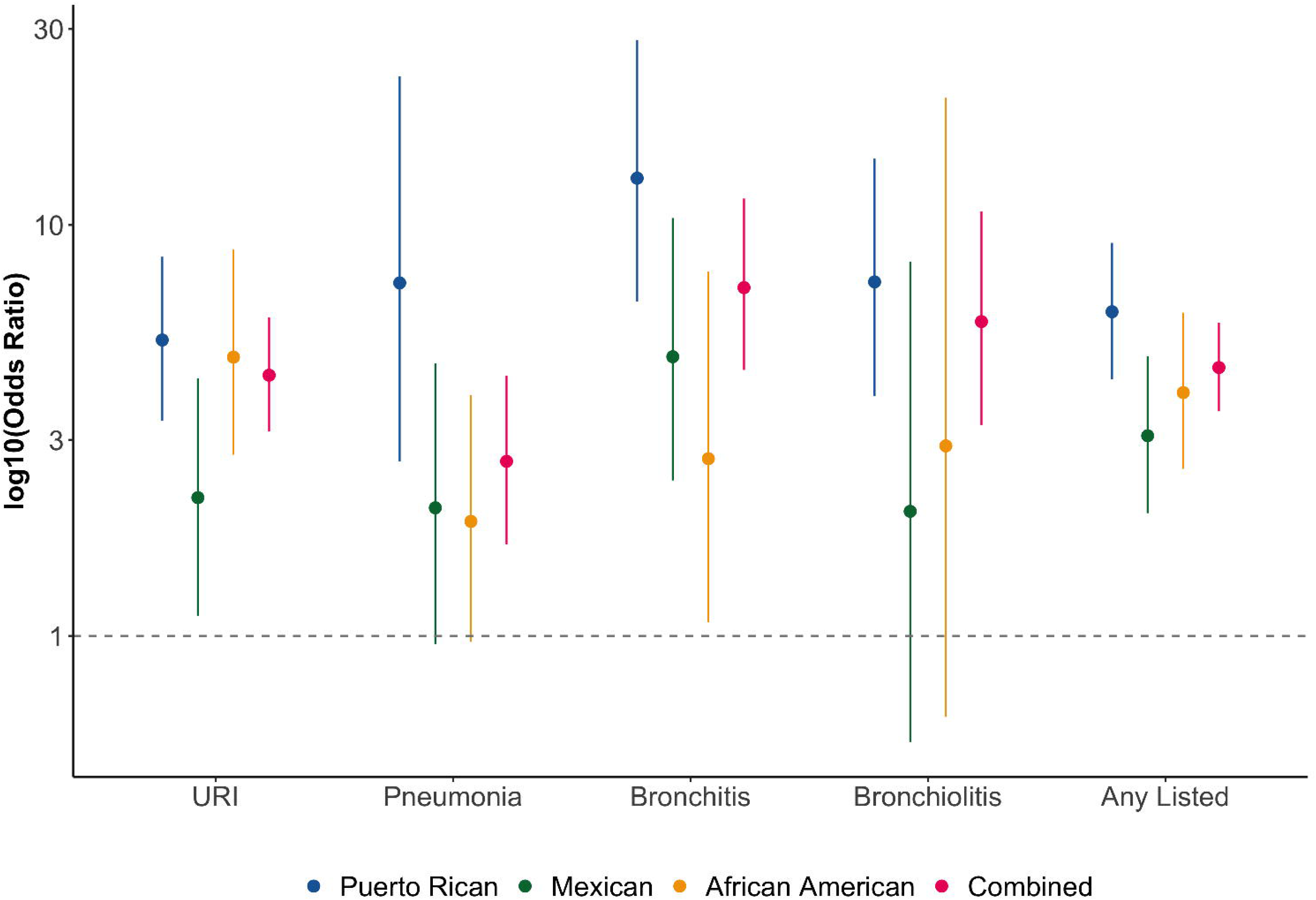
Odds ratios plotted on a log scale of the association between early-life respiratory illnesses before the age of two and asthma diagnosis after the age of two across our three main populations and combined.

To test statistically significant differences in odds ratios between racial/ethnic groups, we performed a Z-test.[29] We found that there were significant differences (p < 0.05) between Puerto Ricans and Mexican Americans for URI and Any Listed, and between Puerto Ricans and African Americans for bronchitis and pneumonia (**Table A in S2 File**).

In a separate analysis, we performed these associations including those individuals diagnosed with asthma before the age of two (**Table B in S2 File**). Our results showed that all early-life respiratory illnesses were significantly associated (p < 0.05) with asthma diagnosis across our three populations and combined, with the exception of RSV in Mexican Americans (**Table C in S2 File**). Additionally, Puerto Ricans had a significantly higher risk of asthma after respiratory infection than Mexican Americans for every respiratory infection tested (**Table D in S2 File**).

We additionally assessed the differential risk of asthma after an early-life respiratory illness in Puerto Ricans living in Puerto Rico (Islanders) versus those living in the mainland United States (Mainlanders); descriptive characteristics for these populations can be found in **Table E in S2 File**. Our data suggest a high asthma risk following early-life respiratory illness in Islanders relative to Mainlanders (**Table F in S2 File**) though due to the small number of Mainlanders we were underpowered to detect statistically significant differences between these groups.

## Discussion

Overall, we found that early-life respiratory illnesses such as URI, pneumonia, bronchitis, and bronchiolitis/RSV are significantly associated with asthma diagnosis after the age of two regardless of race/ethnicity. However, Puerto Ricans displayed the strongest associations.

### Early-life respiratory illnesses increase risk for physician-diagnosed childhood asthma

The association between early-life respiratory illnesses and asthma has been well documented.[8-18] Yet, little is known about racial/ethnic differences that underlie the associations between early-life respiratory illnesses and the development of asthma later on in childhood.

Previous research on the association between early-life respiratory infection and the development of asthma has yielded conflicting results. Having a wheezing illness due to RSV or HRV infection in early life has previously shown to be associated with a 2.6- and 9.8-fold increase, respectively, in asthma risk by age six in a mostly white population.[12] Additionally, a recent study found that the severity of the RSV infection was found to be strongly associated with childhood wheezing at age five in a mostly white population.[11] However, the vast majority of children infected by respiratory viruses like RSV do not go on to develop respiratory illnesses like recurrent wheezing and asthma.[28] Currently, it is unclear why only a minority of children develop asthma after exposure to an early-life respiratory illness. One plausible explanation is that these respiratory illnesses alter the airway in early life, which leads to asthma later on in childhood; previous studies have shown that in adults with asthma, the airway epithelium is altered in a heterogeneous manner.[30] Another possibility is that children who are already genetically or environmentally prone to asthma present with early-life respiratory illness as an early manifestation of asthma. Indeed, previous studies have shown asymptomatic carriage of RSV and rhinovirus in children,[31-33] suggesting a spectrum of disease which may be affected by underlying asthma predisposition.[34, 35] Our results show that asthma risk increases with early-life respiratory illnesses in all populations studied, but that the increase is most dramatic in the Puerto Rican population. These results support previous studies[35-37] that early-life respiratory illnesses are significantly associated with the development of asthma and asthma-related outcomes while additionally adding that these associations may vary by region or population.

### Puerto Ricans are uniquely burdened compared to other groups

Most children who develop respiratory viral infections in early life experience minor illness, but some develop much more severe illnesses that involve lower respiratory symptoms like wheezing.[9, 10, 31] These more severe wheezing illnesses at an early stage in life are associated with a high risk for recurrent wheezing and asthma later in childhood,[12, 13] particularly if these wheezing illnesses were caused by RSV or HRV.[8, 11, 15-18] Puerto Rican children are especially at risk due to the year-round seasonality of RSV infections.[22, 23] In fact, 1,406 cases of bronchiolitis/RSV were reported in children with a mean age of seven months from six Puerto Rican hospitals over a period of nine months.[34] Our results showed that not only is the prevalence of bronchiolitis/RSV infection highest in Puerto Rican children in our study population, but that the risk for asthma among children who have experienced a bronchiolitis/RSV infection in early life is the highest for Puerto Rican Islanders. These observations suggest that the burden of childhood asthma after bronchiolitis/RSV is particularly high Puerto Rican children.

In our analysis of children diagnosed with asthma after two years of age, some confidence intervals were too wide to make definitive conclusions regarding the relative risk of asthma after infection in different racial/ethnic groups. However, when we included those children diagnosed with asthma before two years of age, the increased sample size allowed us to better delineate the association between different respiratory infections and asthma development as stratified by racial/ethnic group. Interestingly, although Puerto Ricans and Mexican Americans are both considered Hispanic/Latino for pulmonary function testing and other health metrics, these populations had widely divergent risks of asthma after early-life respiratory infections. Puerto Ricans consistently had higher risk of asthma after all respiratory infections studied. Whether the association between early-life respiratory infection and increased asthma risk is causal of the alarmingly high rate of asthma in Puerto Ricans requires further study.

While we are not able to draw definitive conclusions about the relative risk of asthma after respiratory infection in Puerto Rican Islanders versus Mainlanders due to the limited number of mainland US Puerto Rican patients studied, we find it interesting that Islander Puerto Ricans tended to have a lower risk of asthma after respiratory infections relative to Islanders. This may suggest an environmental effect about living in Puerto Rico that affects asthma risk. Further studies on Puerto Ricans living on the island and in the mainland US can help clarify this observation.

### Strengths and limitations

Our study may have been limited because the majority (71%) of Puerto Ricans in the GALA study were diagnosed with asthma before the age of two. In contrast, only 35% of Mexican Americans, and 50% of African Americans were diagnosed with asthma before two years old. Our secondary analysis including children diagnosed with asthma before two years of age addressed these differences. Additionally, our exposure measurements were determined retrospectively, which may introduce biases in the data. This study was underpowered to detect any significant associations of early-life respiratory illnesses on asthma in Puerto Rican Mainlanders versus Puerto Rican Islanders, and we were not able to determine whether the year-round RSV prevalence in Puerto Rico modified the effect of the association between respiratory illnesses and asthma. However, our study outlines that there are clear population-specific differences in asthma susceptibility across a variety of respiratory illnesses using a large cohort of minority children from three distinct populations. Our study also benefits from the wide range of clinical, social, and genetic data available on these patients.

## Conclusion

Early-life respiratory illnesses such as RSV, which is highly prevalent in Puerto Ricans, have previously been associated with asthma. Our results indicate that early-life respiratory infections are particularly associated with the later development of asthma among Puerto Rican children compared to other populations.

## Acknowledgements

The authors acknowledge the families and patients for their participation and thank the numerous health care providers and community clinics for their support and participation in GALA II / SAGE. In particular, the authors thank study coordinator Sandra Salazar; the recruiters who obtained the data: Duanny Alva, MD, Gaby Ayala-Rodríguez, Lisa Caine, Elizabeth Castellanos, Jaime Colón, Denise DeJesus, Blanca López, Brenda López, MD, Louis Martos, Vivian Medina, Juana Olivo, Mario Peralta, Esther Pomares, MD, Jihan Quraishi, Johanna Rodríguez, Shahdad Saeedi, Dean Soto, Ana Taveras.

## Supplemental Information

**S1 File. Derivation of variables**. Text A-B.

**S2 File. Supplemental tables**. Tables A-F.

